# Anatomic and Physiologic Scoring is Associated with Mortality and Morbidity Following Operative Intervention for Adult Congenital Heart Disease Patients

**DOI:** 10.64898/2026.07.14.26358111

**Authors:** Brenda La, Jonathan S. Taylor-Fishwick, Morgan MacBeth, Ryo Sakuma, Nicholas Holzemer, Roni Jacobsen, Matthew Stone, Megan SooHoo

## Abstract

**BACKGROUND:** The revised 2018 AHA/ACC guidelines introduced the Adult Congenital Heart Disease Anatomic and Physiologic classification (ACHD-AP) to better categorize disease severity and prognosis in the ACHD population. The ACHD-AP has not been rigorously studied as a perioperative prediction tool.

**OBJECTIVE:** We aimed to assess the accuracy of the ACHD-AP classification in predicting perioperative morbidity and mortality.

**METHODS:** This retrospective cohort study included 295 ACHD patients at a single academic institution between 2018 to 2022. Patients were identified by the STS congenital surgery registry and had undergone a congenital surgical procedure. The primary outcome was overall mortality. Secondary outcomes included short-term post-operative morbidity and comparison of the ACHD-AP score to other existing surgical mortality risk scores. Kaplan-Meier and area under the curve (AUC) of Receiver Operating Characteristic curves were used to evaluate mortality. Logistic regression was used to compare short-term morbidity.

**RESULTS:** A total of 295 patients were included with a median age of 30 years (interquartile range 21-41 years) and 52% were female. There was a total of 14 deaths with 5 (2%) early post-operative deaths and 9 (3%) long-term deaths. By increasing anatomy complexity, overall mortality was 0%, 4% (n=10), and 8% (n=4), respectively. By increasing physiologic severity, overall mortality was 0%, 3% (n=2), 4% (n=7), and 14% (n=5). Moderate and complex anatomy trended towards increased mortality but were not statistically significant (p-value > 0.9). More severe physiology scores predicted increased mortality (p-value = 0.02). Higher physiologic or anatomic complexity scores were associated with longer post-operative length of stay (>5 days). The ACHD-AP AUC was 0.711 for mortality, which was comparable to the Adult Congenital Heart Surgery (ACHS) score (AUC 0.798) and better than the PEACH score (AUC 0.575).

**CONCLUSION:** The ACHD-AP score revealed comparable or better predictive power to existing risk models. Worsening physiologic and anatomy scores were associated with worse post-operative outcomes. Further prospective studies are needed to validate the ACHD-AP score as a prognostic factor for patients undergoing ACHD surgery.

## INTRODUCTION

Survival among adults with congenital heart disease (ACHD) has improved significantly due to advances in prenatal diagnosis, surgical interventions, post-operative care, and long-term care [1, 2]. As a result, approximately 97% of children born with congenital heart disease are expected to reach adulthood, creating a growing population with unique and complex healthcare needs [3, 4]. Many of these patients experience progression of their native disease or long-term sequelae related to prior interventions, often necessitating repeat surgical intervention [5]. As the ACHD population continues to expand, accurate risk stratification tools are increasingly important for perioperative planning, prognostication, and patient counseling.

Traditionally, the Bethesda classification for ACHD categorized disease severity by an anatomic scoring system based on the complexity of the heart defect [2]. The 2018 revised AHA/ACC guidelines introduced the ACHD Anatomic and Physiological (ACHD-AP) classification system that incorporates the aforementioned anatomy scoring alongside new physiological variables [6]. Half of the classification comprises the anatomic class, which categorizes congenital defects by complexities - simple (I), moderate (II), or complex (III). The second component is the physiologic stage, assigned A to D with assessment of multiple physiologic variables including arrhythmia, New York Heart Association classification (NYHA) [7], end organ function, and hypoxemia/cyanosis . These variables were selected based on expert opinion and data existing suggesting their importance in prognosis, management, or quality of life. This provides a total of 12 classifications, based on the highest anatomic class and physiologic staging.

Although ACHD-AP is widely used in clinical practice to categorize disease severity and monitoring disease progression [8-10], its performance as a perioperative risk prediction tool remains poorly defined. The updated 2025 ACC/AHA guidelines recommend ACHD patients with complex underlying cardiac anatomy and physiology receive periprocedural care at a center with ACHD expertise [11]. However, it remains challenging to assess the risk associated with invasive cardiac procedures in patients with ACHD. Existing ACHD surgical mortality risk scores, such as Society of Thoracic Surgery-European Association for Cardio-Thoracic Surgery (STAT) [12], Adult Congenital Heart Surgery (ACHS) [13] and the Perioperative ACHD (PEACH) [14], provide valuable prognostic information but do not incorporate the nuanced physiologic domains emphasized in the ACHD-AP framework.

This study aimed to evaluate the ability of the ACHD-AP classification to predict perioperative morbidity and mortality in adults with congenital heart disease undergoing cardiac surgery and to compare its predictive performance with established ACHD surgical risk scores.

## METHODS

A retrospective cohort study was conducted of adults with congenital heart disease who underwent congenital cardiac surgery at a single academic institution between 2018 and 2022. Patients were identified through the Society of Thoracic Surgeons (STS) congenital surgery registry. Demographic characteristics, ACHD-AP classifications, operative details, and perioperative outcomes were obtained through review of the electronic medical record.

Additional surgical mortality risk scores were calculated for the same cohort using established models, including the Adult Congenital Heart Surgery (ACHS), Society of Thoracic Surgeons-European Association for Cardio-Thoracic Surgery (STAT), and Peri-Operative Risk Assessment in Adult Congenital Heart Disease (PEACH) mortality scores. The primary outcome was all-cause mortality, categorized as short-term (≤ 30 days following the operation) and long-term (>30 days following the operation). Secondary outcomes included post-operative morbidity and comparative assessment of the predictive performance of the ACHD-AP classification relative to existing surgical mortality risk models. Post-operative morbidity outcomes included stroke, post-operative arrhythmia, significant bleeding, hospital readmission, and post-operative length of stay. The majority of missing data were predominantly due to the nature of a retrospective study based on electronic health record review and inconsistent record keeping. Ultimately, these were inconsequential to the analysis of our primary and secondary end points.

All analysis was performed in R version 4.4.3. Statistical significance was set at p < 0.05. Predictive performance of the risk scores for mortality were assessed using area under the curve (AUC) of Receiver Operating Curves (ROC) and Kaplan-Meier survival analysis. Logistic regression models were used to estimate mortality using anatomic and physiologic components independently and combined. Logistic regression was also used to compare short-term morbidity. Fisher exact tests were used to assess differences across groups. Logistic regression models were built to estimate mortality and the combined outcome using anatomic and physiologic components independently and together.

## RESULTS

The ACHD-AP classification was determined for 310 patients. A total of 295 patients were included with a median age of 30 years (interquartile range 21-41 years) and 52% identified as female (**Table 1**). There were 4 patients that were not included in the analysis due to cardiac pathology that could not be defined by the standardized ACHD-AP classification. There were an additional 11 patients who were not included due to duplicate records (the first operation was included in the analysis) thus resulting in 295 final patients. According to the ACHD-AP classification, most patients were of moderate anatomic complexity (n=225, 76%), categorized as class II. The number of patients in physiological stages A, B, C, and D were 14 (4.7%), 64 21.7%), 182 (61.7%), and 35 (11.9%) respectively. There were 6 total patients (2%) categorized in the most severe combined anatomic and physiologic classification of IIID. The most common primary procedures were pulmonic valve replacement (n=30, 10%), anomalous origin of coronary artery repair (n=23, 7.5%), and device implantation (n=23, 7.5%) (**S1**). There was a total of 14 deaths (5%) with 5 (2%) short-term deaths and 9 (3%) long-term deaths. There was no statistical difference between the mean age of those who died compared to those who survived (**Table 2**). The most common causes of death were right ventricular failure, cardiac arrest, and cardiogenic shock (**S2**). Most of these procedures were categorized as elective except for two emergent procedures. Mitral valve repair had the strongest association with mortality.

**Table 1:** Demographics.

| **Age** | **Number** |
| --- | --- |
| Mean (SD) | 32.9 (13.8) |
| Median [Min, Max] | 30.0 [18, 74] |
| **Gender** |  |
| Female | 154 (52.2%) |
| Male | 141 (47.8%) |
| **BMI** |  |
| Mean (SD) | 26.6 (6.0) |
| **Bypass Time (min)** |  |
| Mean (SD) | 167 (90.2) |
| Missing | 43 (14.6%) |
| **Cross clamp Time (min)** |  |
| Mean (SD) | 94.7 (80.4) |
| Median [Min, Max] | 74.0 [0, 333] |
| Missing | 42 (14.2%) |
| **NYHA score** |  |
| Class I | 96 (32.5%) |
| Class II | 131 (44.4%) |
| Class III | 51 (17.3%) |
| Class IV | 17 (5.8%) |
| **ACHD Anatomy Score** |  |
| I (simple) | 20 (6.6%) |
| II (moderate) | 225 (76.3%) |
| III (complex) | 50 (16.9%) |
| **ACHD Physiology Score** |  |
| A | 14 (4.7%) |
| B | 64 (21.7%) |
| C | 182 (61.7%) |
| D | 35 (11.9%) |
| **AP Score** |  |
| I (simple) A | 3 (1.0%) |
| I (simple)B | 8 (2.7%) |
| I (simple)C | 7 (2.4%) |
| I (simple)D | 2 (0.7%) |
| II (moderate)A | 11 (3.7%) |
| II (moderate)B | 52 (17.6%) |
| II (moderate)C | 135 (45.8%) |
| II (moderate)D | 27 (9.2%) |
| III (complex)B | 4 (1.4%) |
| III (complex)C | 40 (13.6%) |
| III (complex)D | 6 (2.0%) |

**Table 2:** Event and mortality.

| Event | Total<br>(n = 295) | No Mortality<br>(n = 281) | Mortality<br>(n = 14) | p-value |
| --- | --- | --- | --- | --- |
| **Mortality** |  |  |  |  |
| Short-Term Mortality (<=30 days) |  |  | 5 (35.7%) | <0.001 |
| Long-Term Mortality (>30 days) |  |  | 9 (64.3%) | <0.001 |
| Mean age [IQR] | 33 [21 – 41] | 30 [21 – 41] | 34 [29 – 48] | 0.13 |
| BMI (kg/m<sup>2</sup>) | 27 | 25 | 24 | 0.2 |
| **Post-operative Outcomes** |  |  |  |  |
| Post-op length of stay (avg days) | 8 | 5 | 8 | 0.5 |
| Post-op arrhythmia | 78 (26.4%) | 77 (27.4%) | 1 (7.1%) | 0.2 |
| Post-op Stroke | 6 (2.0%) | 5 (1.8%) | 1 (7.1%) | 0.2 |
| Endocarditis | 30 (10.2%) | 29 (10.3%) | 1 (7.1%) | >0.9 |
| 30-day readmission | 28 (9.5%) | 28 (10.0%) | 0 (0%) | >0.9 |
| Bypass time (avg min) | 171 | 155 | 311 | **0.003** |
| Reoperation for bleeding | 8 (2.7%) | 6 (2.1%) | 2 (14.3%) | **0.044** |
| Cross clamp time (avg min) | 96.2 | 71 | 123 | 0.2 |
| **ACHD-AP Scores** |  |  |  |  |
| Anatomy Score |  |  |  | >0.9 |
| I (simple) | 20 (6.8%) | 20 (71.2%) | 0 (0%) |  |
| II (moderate) | 224 (75.9%) | 215 (76.5%) | 9 (64.3%) |  |
| III (complex) | 50 (16.8%) | 46 (16.4%) | 4 (28.6%) |  |
| Physiology Score |  |  |  | 0.02 |
| A | 14 (4.7%) | 14 (5.0%) | 0 (0%) |  |
| B | 64 (21.7%) | 62 (22.1%) | 2 (14.3%) |  |
| C | 181 (61.4%) | 174 (61.9%) | 7 (50%) |  |
| D | 35 (11.9%) | 31 (11.0%) | 4 (28.6%) |  |
| AP Score |  |  |  | 0.5 |
| IA | 3 (1.0%) | 3 (1.1%) | 0 (0%) |  |
| IB | 8 (2.7%) | 8 (2.8%) | 0 (0%) |  |
| IC | 7 (2.4%) | 7 (2.5%) | 0 (0%) |  |
| ID | 2 (0.7%) | 2 (0.7%) | 0 (0%) |  |
| IIA | 11 (3.7%) | 11 (3.9%) | 0 (0%) |  |
| IIB | 52 (17.6%) | 50 (17.8%) | 2 (14.3%) |  |
| IIC | 134 (45.4%) | 130 (46.3%) | 4 (28.6%) |  |
| IID | 27 (9.2%) | 24 (8.5%) | 3 (21.4%) |  |
| IIIB | 4 (1.4%) | 4 (1.4%) | 0 (0%) |  |
| IIIC | 40 (13.6%) | 37 (13.2%) | 3 (21.4%) |  |
| IIID | 6 (2.0%) | 5 (1.2%) | 1 (7.1%) |  |
| **Risk Tools** |  |  |  |  |
| NYHA Score (III-IV) | 68 | 6 | 62 | 0.065 |
| ACHS Score >= 1 | 31 | 27 | 4 | **0.041** |
| STAT category >= 3 | 51 | 46 | 5 | 0.055 |

By increasing isolated anatomical complexity, overall mortality was 0% (n=0), 5% (n=11), and 4% (n=3), respectively (p>0.9). By increasing isolated physiologic severity, overall mortality was 0% (n=0), 3% (n=2), 4% (n=7), and 14% (n=5) with a p of 0.02 (**Table 3**). There was no observed mortality for anatomy class I or physiology stage A. Moderate and complex anatomy trended towards increased mortality; however, the association was not statistically significant (p>0.9). Worsening physiology score was only significant among anatomy class II patients as most patients met criteria for this anatomic classification (**Table 4**).

**Table 3.** Mortality in Independent ACHD-AP Scores.

| Primary Outcome in Independent ACHD-AP Components |  |  |  |  |  |
| --- | --- | --- | --- | --- | --- |
| Anatomy Class | I<br>n = 0/20 | II<br>n = 9/224 | III<br>n = 4/50 |  | p-value |
| Mortality | 0% | 4% | 8% |  | >0.9 |
| Physiology Stage | A<br>n = 0/14 | B<br>n = 2/64 | C<br>n = 7/181 | D<br>n = 5/35 | p-value |
| Mortality | 0% | 3% | 4% | 14% | **0.02** |

**Table 4.** Mortality Across ACHD-AP Scores.

| Primary Outcome Across ACHD-AP Scores |  |  |  |  |  |  |  |  |  |  |  |  |  |  |  |
| --- | --- | --- | --- | --- | --- | --- | --- | --- | --- | --- | --- | --- | --- | --- | --- |
| Anatomy | I |  |  |  |  | II |  |  |  |  | III |  |  |  |  |
| Physiology | A<br>n=3 | B<br>n=8 | C<br>n=7 | D<br>n=2 | P | A<br>n=11 | B<br>n=52 | C<br>n=139 | D<br>n=30 | P* | A<br>n=0 | B<br>n=4 | C<br>n=43 | D<br>n=6 | P |
| Mortality | 0<br>0% | 0<br>0% | 0<br>0% | 0<br>0% | >0.9 | 0<br>0% | 3<br>5.8% | 5<br>3.6% | 6<br>20% | **0.017** | 0<br>0% | 0<br>0% | 2<br>4.7% | 1<br>17% | 0.5 |

Although the Kaplan Meier survival analysis demonstrates distinct survival curves for patients categorized with AP scores IID, IIIC, and IIID (**Figure 1**), this was not statistically significant for the follow-up time considered.

**Figure 1.**
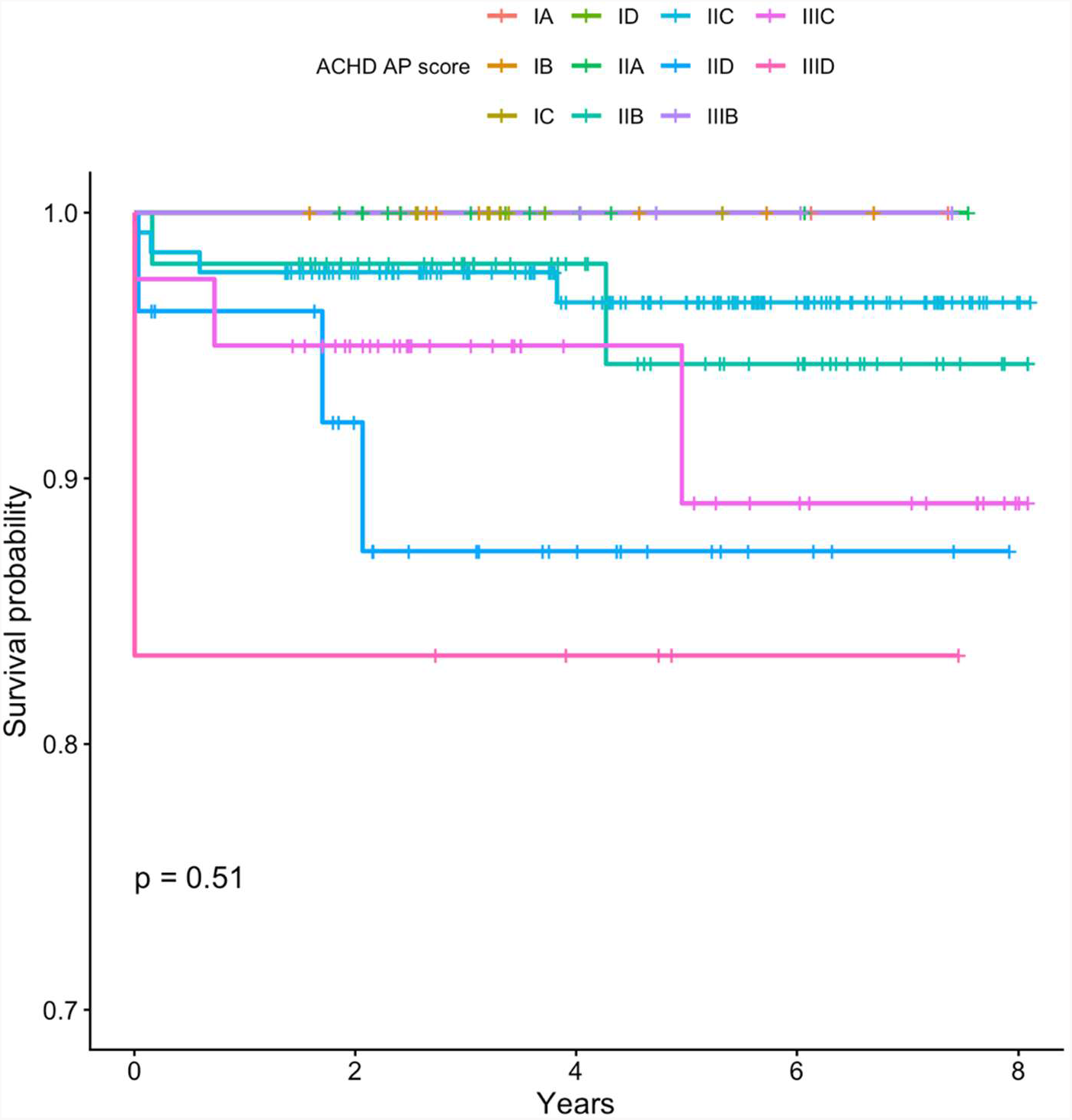
Kaplan Meier Survival Curve for ACHD-AP Scores

Secondary outcomes of post-operative stroke, arrhythmia, bleeding, 30-day readmission, and length of stay (LOS) were also assessed using ACHD-AP classifications (**Table 5**). When compared within the same category of anatomic class, higher physiologic complexity was associated with higher occurrence of secondary outcomes. This was statistically significant for the anatomic classification II (p 0.001), however was not statistically significant for anatomy I and III. Higher physiologic and anatomic complexity scores were associated with longer post-operative length of stay and 30-day readmission rate, both were statistically significant (p<0.05). Other post-operative complications demonstrated a similar trend but were less consistent across classes.

**Table 5:** Post-operative Morbidity Across Physiology Scores.

| Secondary Outcomes Across ACHD Physiology Scores |  |  |  |  |  |
| --- | --- | --- | --- | --- | --- |
| Post-Op Morbidity | A<br>n = 14 | B<br>n = 64 | C<br>n = 182 | D<br>n = 35 | p-value |
| Stroke | 0% | 2% | 2% | 3% | 0.9 |
| Arrhythmia | 21% | 16% | 29% | 37% | 0.08 |
| Bleeding | 0% | 2% | 2% | 9% | 0.2 |
| 30-day readmission | 14% | 0% | 13% | 13% | **0.004** |
| Length of Stay (days) | 4 | 4 | 6 | 9 | **<0.001** |

**Table 6.** Post-operative Morbidity Across Anatomy Scores.

| Secondary Outcomes Across ACHD Physiology Scores |  |  |  |  |
| --- | --- | --- | --- | --- |
| Post-Op Morbidity | I<br>n = 20 | II<br>n = 225 | III<br>n = 50 | p-value |
| Stroke | 0% | 2% | 2% | >0.9 |
| Arrhythmia | 10% | 30% | 20% | 0.08 |
| Bleeding | 0% | 3% | 2% | >0.9 |
| 30-day readmission | 0% | 10% | 14% | 0.2 |
| Length of Stay (days) | 4 | 5 | 7 | **0.04** |

AUC analysis was used to compare the discriminative power of the ACHD-AP score for overall mortality to various other prognostic scoring systems. The AUC for overall mortality for ACHD-AP, PEACH, and ACHS was 0.711, 0.575, 0.798, respectively **(Figures 2 – 4)**.

**Figure 2.**
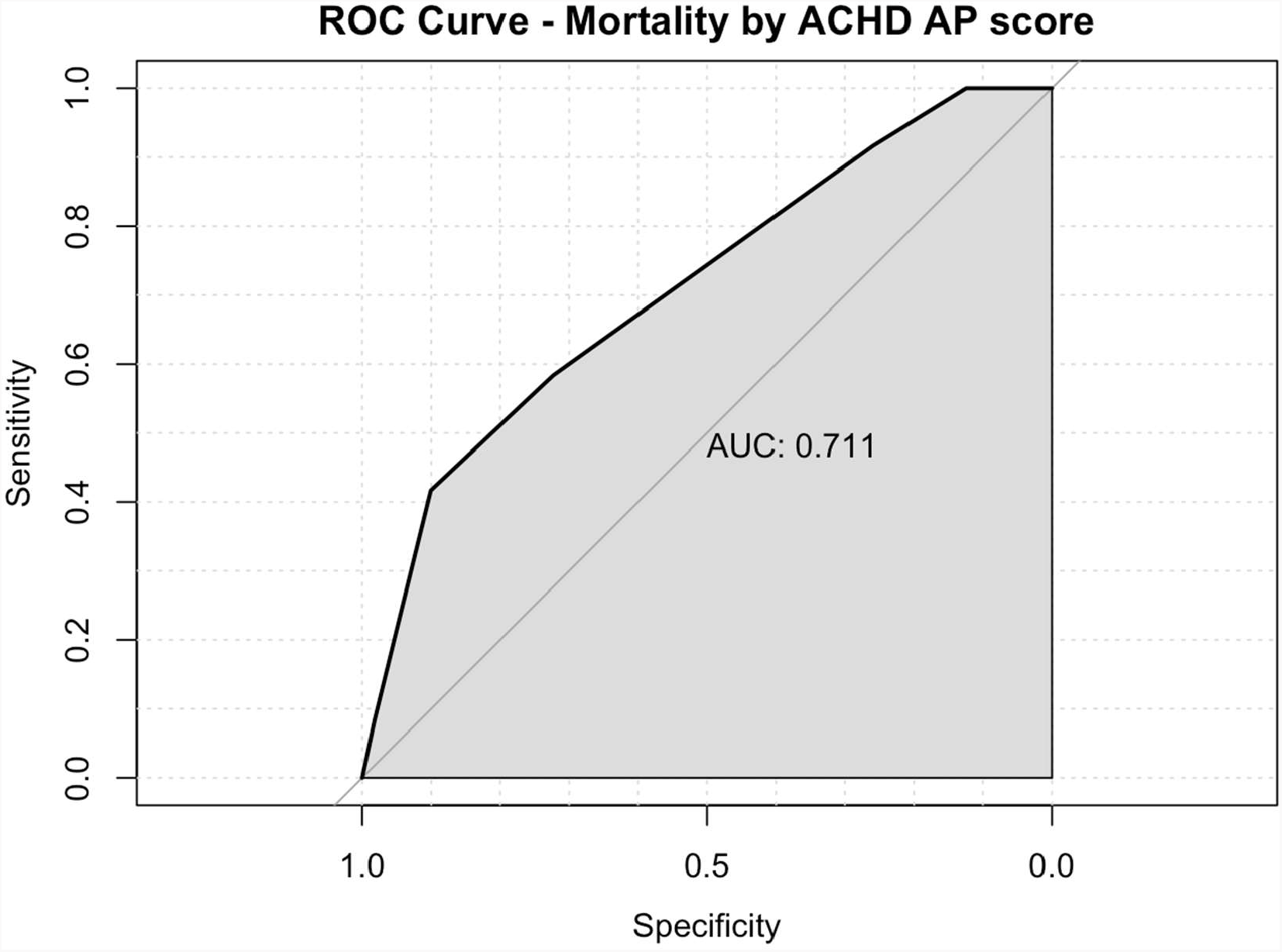
AUC ACHD-AP Score

**Figure 3.**
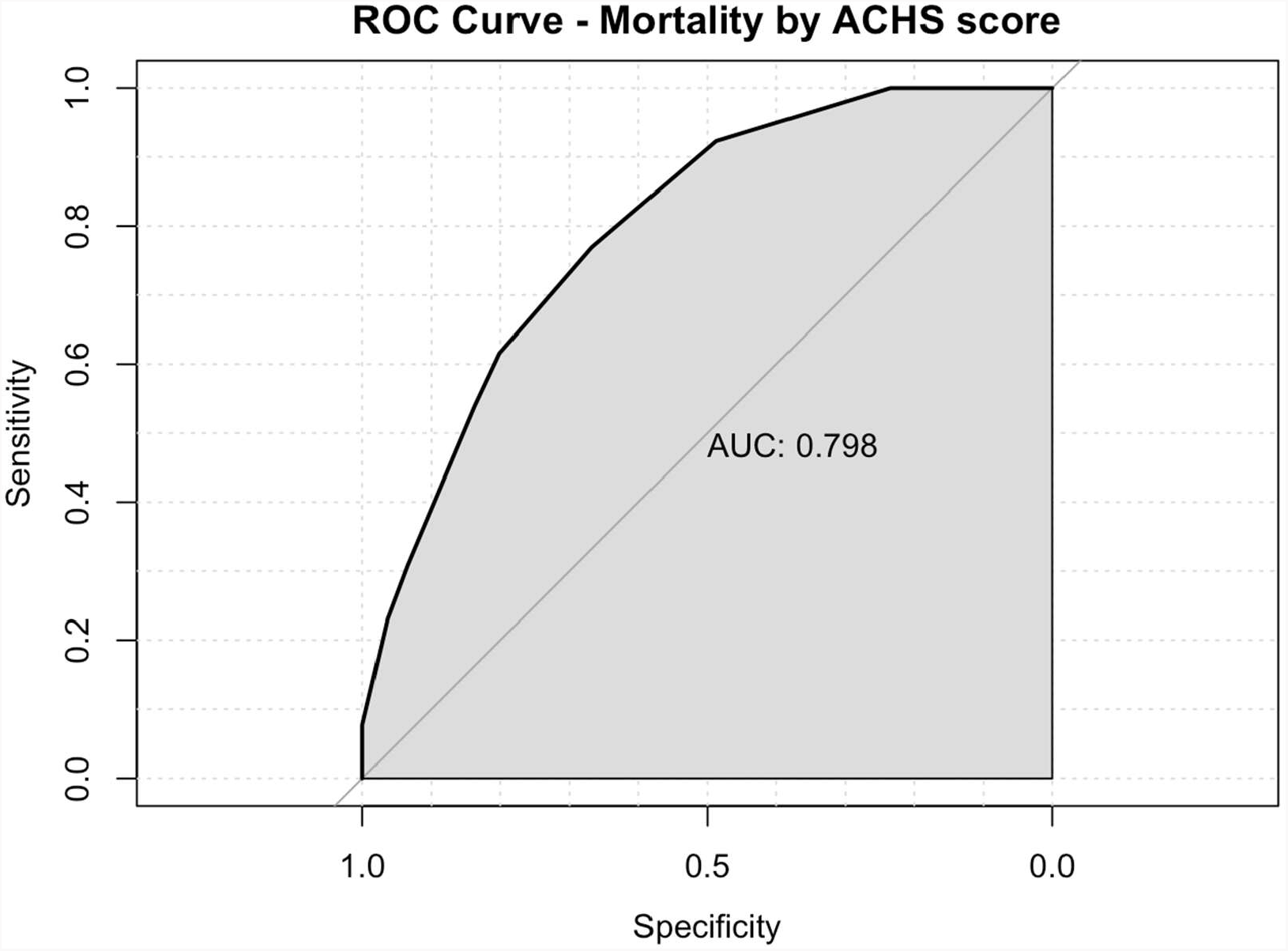
AUC ACHS Score

**Figure 4.**
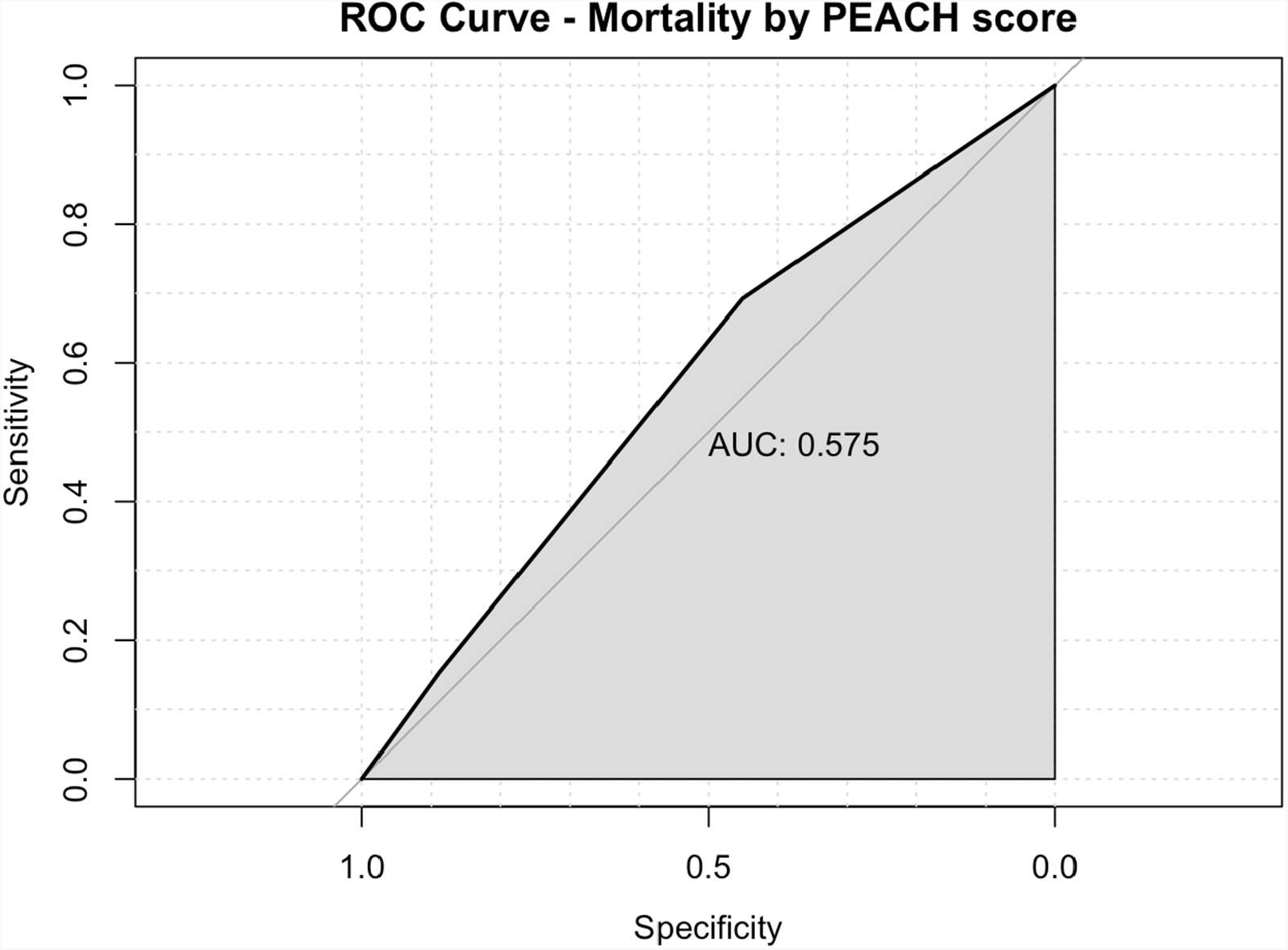
AUC PEACH Score

## DISCUSSION

In this single-center retrospective cohort of adults with congenital heart disease undergoing cardiac surgery, the ACHD-AP classification was associated with post-operative outcomes, more strongly driven by physiologic stage than anatomic complexity. Increasing physiologic severity demonstrated a significant association with mortality, whereas anatomic complexity alone showed limited discriminatory value. Overall, the ACHD-AP score demonstrated moderate predictive ability for mortality and performance comparable to the established ACHS risk model and better than the PEACH score. In addition, higher physiologic burden was associated with increased post-operative morbidity, including prolonged length of stay, supporting the clinical relevance of physiologic assessment in risk stratification.

Worsening physiologic stage within the ACHD-AP classification was significantly associated with increased perioperative mortality, with mortality progressively increasing from 0% in stage A to 11% in stage D (p=0.02). In contrast, anatomic complexity alone was not significantly associated with mortality, despite a numerical trend toward higher mortality among patients with moderate and complex anatomy. While this may partially reflect limited sample size, particularly within the complex anatomy group, these findings also suggest that the original congenital heart defect alone may not adequately capture operative risk in the ACHD population, especially those undergoing repeat intervention. Patients with similar underlying anatomic lesions may have markedly different physiologic burden depending on several factors including ventricular function, arrhythmia burden, end-organ dysfunction, exercise intolerance, or prior surgical sequelae - all of which may substantially influence perioperative vulnerability [15].

Importantly, worsening physiologic stage remained associated with mortality even within the same anatomic classification, further supporting the incremental prognostic value of physiologic assessment beyond anatomy alone. The stronger association between physiologic stage and mortality is clinically intuitive, as physiologic classification likely reflects a patient’s cumulative disease burden and physiologic reserve at the time of surgery rather than anatomy alone. Advances in congenital cardiac surgery and longitudinal ACHD care have improved survival across a broad range of anatomic complexity [2, 16], resulting in a growing population of adults whose perioperative risk may be driven less by their original diagnosis and more by the downstream consequences of chronic cardiovascular disease. These findings are consistent with prior studies demonstrating improved prediction of all-cause mortality when physiologic status is integrated into congenital heart disease classification systems [9].

Beyond mortality, the ACHD-AP classification also demonstrated associations with clinically meaningful post-operative morbidity. Higher physiologic stage was associated with prolonged hospital length of stay and increased incidence of 30-day readmission, with patients classified as physiologic stage D experiencing the longest hospitalizations and highest readmission rates. Importantly, worsening physiologic stage remained associated with increased post-operative morbidity even within the same anatomic classification, particularly among patients with moderate anatomic complexity. These findings further suggest that physiologic status provides valuable incremental information beyond anatomy alone when assessing perioperative risk in ACHD patients and their potential vulnerability to post-operative complications, prolonged recovery, and recurrent hospitalization.

While complications such as stroke or bleeding were less consistently associated with ACHD-AP classification, broader markers of post-operative recovery, including length of stay and readmission, demonstrated significant associations. Collectively, these findings suggest that physiologic staging may help anticipate post-operative complications and recovery trajectory, support perioperative planning and resource allocation by the multidisciplinary care team, and facilitate more informed preoperative counseling for ACHD patients undergoing cardiac surgery.

Existing ACHD surgical risk models such as STAT, ACHS, and PEACH were primarily designed to estimate operative mortality using procedural complexity and registry-derived factors. While these tools provide important perioperative risk estimation, they may not fully capture the longitudinal physiologic burden and long-term sequelae characteristic of the ACHD population. In our study, the ACHD-AP score demonstrated moderate discrimination for mortality, with performance comparable or superior to established risk models. Although ACHS demonstrated slightly higher discriminatory ability, the ACHD-AP framework offers the distinct advantage of incorporating physiologic status in addition to anatomic complexity, potentially providing a more comprehensive assessment of perioperative vulnerability. Importantly, the ACHD-AP classification can be readily applied during routine clinical evaluation without requiring complex procedural calculations or additional scoring tools. As such, it may serve as a practical complementary framework to existing surgical risk models rather than a replacement.

## LIMITATIONS

This study has several limitations. As a retrospective, single-center analysis, generalizability is limited. The number of mortality events was small, reducing statistical power and limiting subgroup analyses. Unequal distribution across ACHD-AP categories further constrained comparisons between groups. Additional limitations include reliance on clinical documentation for classification, potential misclassification, procedural heterogeneity, and unmeasured confounding. The ACHD-AP framework also does not account for prior surgical history or differential weighting of physiologic variables, which may influence operative risk.

Prospective, multicenter studies are needed to validate these findings and further evaluate the predictive performance of the ACHD-AP classification. Future work should assess whether incorporating additional clinical variables, biomarkers, or prior operative history improves model discrimination. Given the dynamic nature of physiologic classification, longitudinal assessment of changes in physiologic stage before and after surgery may further refine its prognostic utility.

## CONCLUSION

In this cohort of adults with congenital heart disease undergoing cardiac surgery, the ACHD-AP classification was associated with perioperative outcomes, with physiologic stage more strongly linked to adverse outcomes than anatomic complexity. The ACHD-AP score demonstrated moderate discrimination for mortality and comparable or superior performance to established surgical risk models. Physiologic severity was the most consistent predictor of morbidity and mortality, including prolonged hospitalization. These findings suggest the clinical utility of the ACHD-AP framework for perioperative risk assessment and patient counseling prior to surgical intervention, particularly through incorporation of physiologic status. Overall, the ACHD-AP classification provides risk stratification in this population and appears to be a potentially useful, accessible, and comprehensive perioperative prognostic tool in ACHD care. Further multi-center studies with larger, more standardized cohorts are needed to validate and refine current risk prediction tools for this unique patient population.

## Data Availability

Data are available from the authors upon reasonable request. Some data may be limited to public sharing due to HIPAA and patient privacy.

## Supplementary Material

**S1:** Table describing causes of mortality.

| **Cause of death** | **N (% of Mortality)** |
| --- | --- |
| RV Failure | 3 (21%) |
| Cardiac arrest | 3 (21%) |
| Cardiogenic shock | 2 (14%) |
| Myocardial ischemia | 1 (7%) |
| Stroke | 1 (7%) |
| Post-op hemorrhage | 1 (7%) |
| Metastatic cancer | 1 (7%) |
| ESRD | 1 (7%) |
| Unknown | 1 (7%) |

**S2:** Table listing occurrences of different cardiac repairs.

**S2:** Table listing occurrences of different cardiac repairs
| **Repair** | **Occurrence (% of total)** |
| --- | --- |
| Pulmonic valve replacement | 30 (10%) |
| Anomalous origin of coronary artery | 23 (7.7%) |
| Device implantation | 23 (7.7%) |
| Aortic aneurysm repair | 22 (7.3%) |
| ASD repair | 16 (5.3%) |
| PAPVC repair | 15 (5.0%) |
| Valvuloplasty | 14 (4.7%) |
| Mitral valve replacement | 13 (4.3%) |
| Aortic root replacement | 12 (4.0%) |
| Tricuspid valve replacement | 12 (4.0%) |
| Heart transplant | 11 (3.7%) |
| Explanation of pacing system | 10 (3.3%) |
| Ross procedure | 10 (3.3%) |
| Conduit reoperation | 9 (3.0%) |
| Aortic valve replacement | 9 (3.0%) |
| Aortic stenosis with repair | 7 (2.3%) |
| Atrial arrhythmia ablation | 6 (2.0%) |
| Removal of transcatheter-delivered device from heart | 6 (2.0%) |
| Systemic venous stenosis repair | 6 (2.0%) |
| VSD repair | 6 (2.0%) |
| Vascular ring repair | 5 (1.7%) |
| Ebstein's Repair | 4 (1.3%) |
| Konno Procedure | 4 (1.3%) |
| PA Reconstruction | 4 (1.3%) |
| RVOT Procedure | 4 (1.3%) |
| Ventricular arrhythmia ablation | 3 (1.0%) |
| Conduit placement RV to PA | 2 (0.7%) |
| Pulmonary venous stenosis repair | 2 (0.7%) |
| PA aneurysm repair | 1 (0.3%) |
| Aortic arch repair | 1 (0.3%) |
| ASD creation/enlargement | 1 (0.3%) |
| Coarctation repair | 1 (0.3%) |
| Coronary artery bypass graft | 1 (0.3%) |
| Intravascular stent removal | 1 (0.3%) |
| Pericardial drainage | 1 (0.3%) |
| Pericardiectomy | 1 (0.3%) |
| Ross-Konno Procedure | 1 (0.3%) |
| Sinus of Valsalva aneurysm | 1 (0.3%) |
| Sternotomy wound drainage | 1 (0.3%) |
| Truncal valve replacement | 1 (0.3%) |

**S3:**
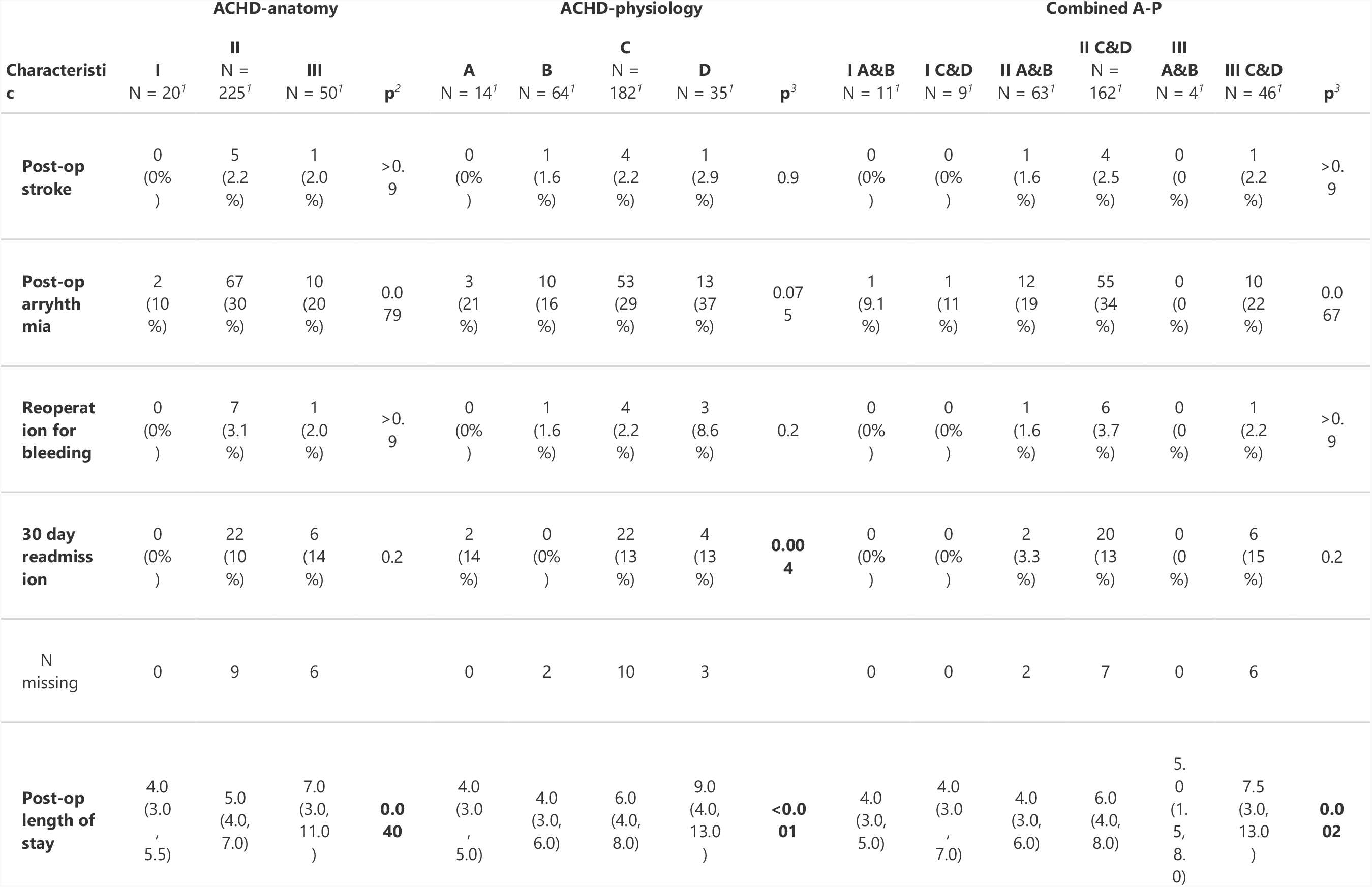
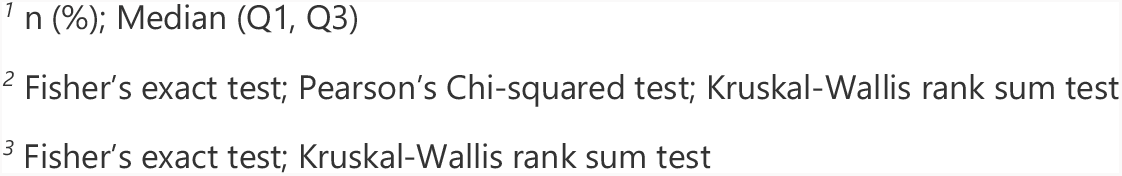
Table of secondary outcomes across ACHD scores.

